# Iron status, thyroid dysfunction and iron deficiency anemia: a two-sample Mendelian randomization study

**DOI:** 10.1101/2023.11.15.23298576

**Authors:** Xianjun Huang, Tianhong Guo, Yuqin Wu, Qi Xu, Junliang Dai, Yuanshuai Huang

**Author notes:** Corresponding Author: Yuanshuai Huang.

## Abstract

**Objective:** Given the clinical association between thyroid dysfunction and iron deficiency anemia (IDA), as well as their shared association with iron status, this study aims to investigate the causal relationship between iron status and thyroid dysfunction, while also examining the risk of IDA in relation to thyroid dysfunction.

**Methods:** A two-sample Mendelian randomization (MR) study was conducted to identify the causal relationship of iron status on thyroid dysfunction, as well as thyroid dysfunction on IDA. Large-scale European population-based GWAS databases were utilized (Genetics of Iron Status consortium, ThyroidOmics consortium, FinnGen consortium, and UK biobank). Inverse variance weighted (IVW) was used as the main analysis. In addition, we used weighted median and MR-Egger to enhance the robustness. Sensitivity analysis was conducted to evaluate the robustness of MR results.

**Results:** The IVW estimates did not reveal any significant causal relationship between serum iron status markers and thyroid dysfunction. However, a significant causal relationship was observed between hypothyroidism and IDA (OR = 1.101, 95% CI = 1.048-1.157, *p* < 0.001). Repeated analyses also demonstrated a similar trend (OR = 1.023, 95% CI = 1.011-1.035, *p* < 0.001). Sensitivity analysis supported that the MR estimates were robust.

**Conclusion:** In our MR study, an upregulation of the hypothyroidism-associated gene was found to be significantly associated with an elevated risk of IDA in the European population. These findings may offer novel therapeutic insights for clinicians managing patients with hypothyroidism, IDA, or their comorbidities.

## Introduction

Anemia is quantitatively defined as a decrease in the number of circulating erythrocytes, or functionally defined as a condition characterized by an insufficient count of red blood cells (oxygen carriers) to meet an individual’s metabolic demands[1]. Anemia can be attributed to various etiologies, encompassing augmented loss or destruction of red blood cells and inadequate production or generation of defective red blood cells[2]. Iron deficiency anemia (IDA) is classified as one of the various types of anemia, with iron deficiency being the predominant etiology[3]. IDA is a form of nutritional anemia resulting from insufficient production of hemoglobin in the body due to iron deficiency[4]. The prevalence of IDA affects 30% of the global population, with a higher incidence observed among children[5]. In 2016, there were over 1.2 billion reported cases of IDA worldwide. IDA ranks among the top five causes of disability globally and stands as the primary cause in low– and middle-income countries; moreover, it is also recognized as the leading cause among women in 35 nations [6].

The dysfunction of the thyroid gland is a disorder that affects the endocrine function of the pituitary-hypothalamic-thyroid axis, resulting from various physiological or pathological factors. This ultimately leads to an imbalance in circulating thyroxine levels, either excessively high or low, thereby failing to meet the body’s normal endocrine and metabolic requirements. It can be broadly categorized into subclinical hyperthyroidism or hypothyroidism (abnormal TSH levels with normal thyroxine levels)[7], and overt hyperthyroidism or hypothyroidism (abnormal thyroxine levels)[8]. Thyroid dysfunction can manifest at any age, including childhood[9], as well as exert detrimental effects on fetal development in pregnant women and obstetric outcomes[10]. The occurrence of thyroid dysfunction is closely associated with aberrant immune function and numerous immune factors[11].

Numerous researchers posit that the iron status of the body exerts an influence on thyroid function. Initial animal experiments have demonstrated that iron deficiency can potentially alter the central regulation of the thyroid axis[12], while hypothyroid rats exhibit reduced gastrointestinal iron absorption, which is restored to normal levels following T3 supplementation[13]. Iron deficiency can also diminish the thyroid hormone production activity of thyroid peroxidase (TPO), an iron-dependent enzyme[14–16]. Immune thyroid diseases frequently coexist with other autoimmune disorders, such as autoimmune gastritis or intestinal conditions[17, 18], which can potentially impact the absorption of essential nutrients like iron. In two studies involving patients with concurrent iron deficiency anemia and subclinical hypothyroidism, the combined administration of iron and T4 demonstrated superior efficacy in improving iron status compared to monotherapy alone[19, 20]. However, some studies have reported no significant correlation between serum thyroid hormone concentration and iron status[21, 22], indicating a different situation. Numerous observational studies have consistently demonstrated a significant association between thyroid function and anemia. In a study conducted by Erdogan et al.[23], the prevalence of anemia was found to be comparable in patients with overt and subclinical hypothyroidism, at 43% and 39%, respectively. Additionally, a prospective study by Chris-Crain et al.[24] revealed that restoration of thyroid function resulted in elevated erythropoietin concentrations among individuals with subclinical thyroid dysfunction. However, a prospective cohort study and systematic review yielded contrasting findings[25], suggesting that subclinical thyroid dysfunction does not exhibit any association with alterations in hemoglobin concentration during follow-up, nor does it serve as an independent risk factor for the development of anemia, changes in Hb concentration among patients with overt thyroid dysfunction lack clinical significance. In summary, the relationship between iron status and thyroid function remains inconclusive, as does the causal association between thyroid function and iron deficiency anemia. Elucidating these causal relationships holds significant implications for enhancing prevention and treatment strategies for both thyroid diseases and IDA.

Due to limited evidence and conflicting conclusions from previous observational studies, a consensus regarding the precise relationship between iron status and thyroid function, as well as between thyroid function and IDA, is currently lacking. Further investigation is warranted to provide more compelling evidence. Mendelian randomization (MR), an emerging methodology for establishing potential causal relationships between exposure factors and outcomes, utilizes specific single nucleotide polymorphisms (SNPs) as instrumental variables (IVs)[26]. This study design is robust against potential confounding or reverse causality due to the random distribution of midrange genes during gametic formation, ensuring its suitability for elucidating causal effects between exposure and outcomes. With the increasing availability of large-scale genome-wide association study (GWAS) data in the public domain, methods that achieve high statistical power become more effective. MR designs have previously been employed to investigate the causal relationship between thyroid function and various outcome features such as liver cancer[27], lung cancer[28], and benign prostate disease[29]. However, these designs have not yet been utilized to examine the impact of iron status on thyroid dysfunction or the effect of thyroid dysfunction on IDA. Therefore, we conducted a two-sample MR study to investigate the causal relationship between biochemical markers of iron status, thyroid dysfunction, and IDA.

## Methods

### Study design

In the present two-sample MR study, we utilized data from two distinct GWAS, one for exposure and another for outcome, to estimate the causal effect of exposure on outcome. In all corresponding original studies, informed consent was obtained from all participants, and since only summary-level statistics were employed in our analysis, no additional ethical approval was required. The initial analysis comprised three two-sample MR analyses. The first MR analysis was conducted using the Genetics of Iron Status consortium (GIS)[30] dataset (n = 23,986), which included biochemical markers of iron status such as serum iron, ferritin, transferrin saturation, and transferrin levels. Additionally, we utilized subclinical hyperthyroidism and hypothyroidism datasets from the ThyroidOmics consortium[31](n = 72,167), as well as overt hyperthyroidism (n = 460,499) and hypothyroidism (n = 410,141) data from the GWAS Catalog. The second MR analysis incorporated the aforementioned data on thyroid dysfunction and IDA (n = 217,202) from the FinnGen consortium. To establish a causal relationship between iron status and IDA, we conducted a third MR analysis using data on the biochemical markers of iron status mentioned above, in conjunction with the FinnGen consortium’s IDA dataset. In order to ensure non-overlapping populations, GWAS data for exposure and outcomes were sourced from distinct databases. To validate the robustness of the significance results of the primary analysis, we conducted a replicated validation using exposure and outcome GWAS data from different databases. Table 1 presents all the GWAS data incorporated in this study.

**Table 1.**
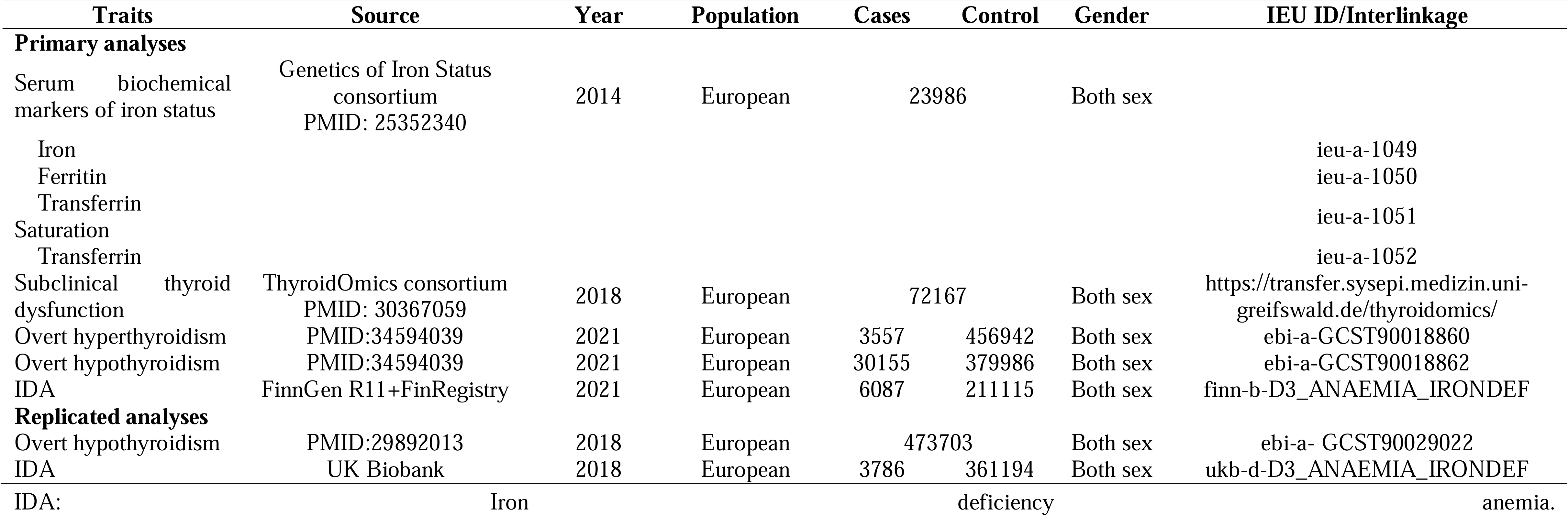
GWAS datasets information overview.

As shown in Figure 1, a two-sample MR study must adhere to three fundamental assumptions. Hypothesis 1 posits that genetic instrumental variables exhibit strong associations with exposure factors. Given the random allocation of SNPs during conception, hypothesis 2 suggests that the genetic instrumental variables for exposure factors remain unaffected by confounding factors. Hypothesis 3 asserts that outcome characteristics are predominantly influenced by the genetic instrumental variables of exposure factors rather than alternative mechanisms[32]. Collectively, hypotheses 2 and 3 are referred to as being independent of pleiotropy[26].

**Fig. 1.**
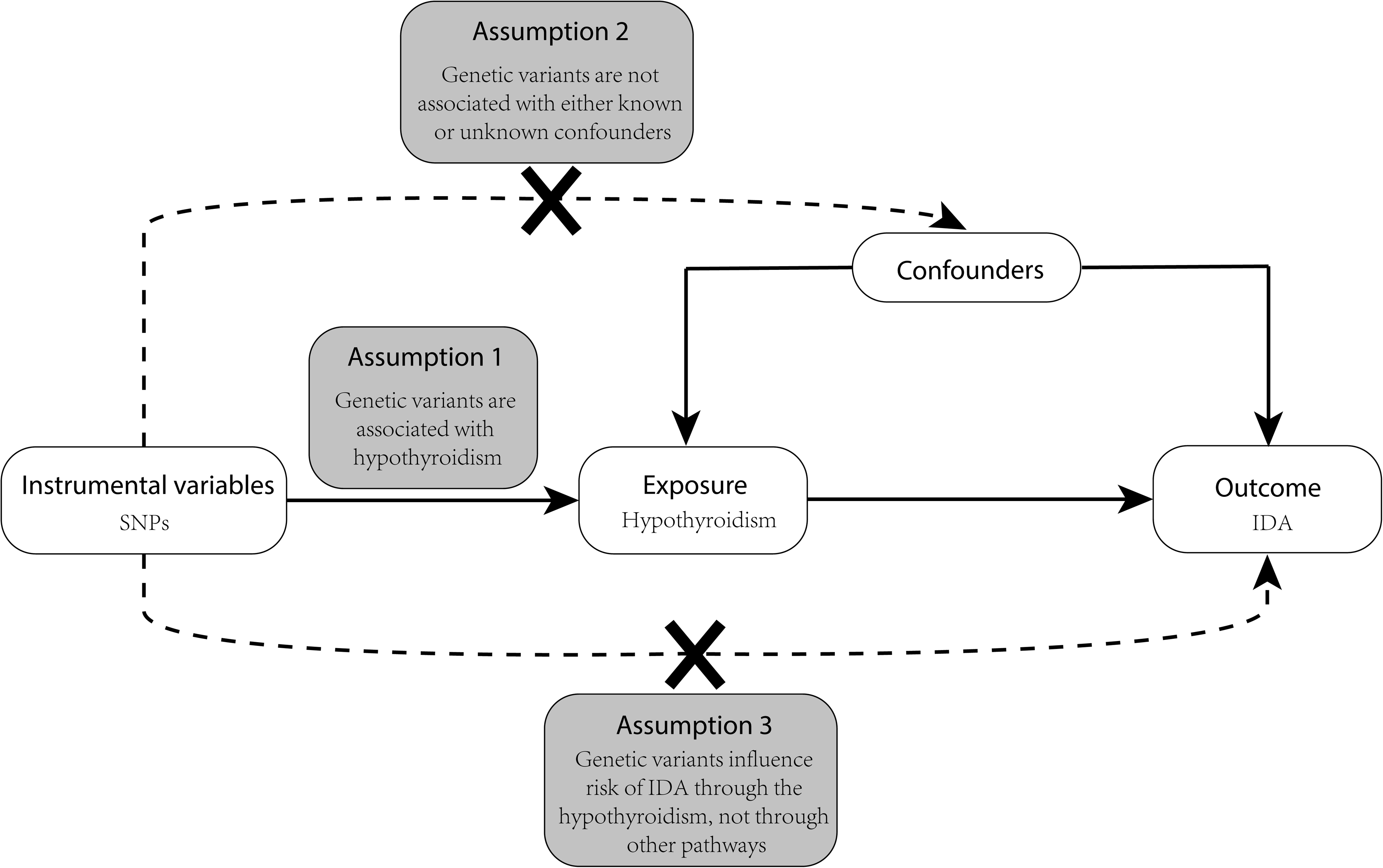
Three core assumptions of Mendelian randomization.

In MR studies, sensitivity analysis is an essential method for evaluating potential bias, encompassing two key considerations: a heterogeneity test and a pleiotropy test. The detection of heterogeneity in IVW methods was performed using Cochran’s Q test, while the presence of horizontal pleiotropy was indicated by an intercept with *p* < 0.05 in MR-Egger regression[33]. Moreover, we employed MR-PRESSO and Radial MR to eliminate SNPs with pleiotropic outliers (*p* < 0.05)[34, 35]. Upon identification of potential outliers, we excluded them from the analysis and conducted a repeated IVW estimate to evaluate the robustness of our findings. To mitigate reverse causality, we employed the Steiger Test[36]. Additionally, a leave-one-out analysis was performed to assess the potential influence or bias of individual SNPs on the MR estimate. As implied by its name, this approach involved iteratively excluding one SNP at a time and re-performing MR analyses, thereby enabling an evaluation of whether the causal estimate was driven by any single SNP.

### Selection of genetic variation SNPs

Typically, genome-wide significant genetic variation (*p* < 5×10^-8^) is employed as a potential tool. To enhance the independence of single SNP, we pruned these tools within a window size of 10,000 kb to mitigate linkage disequilibrium (LD) at a threshold of r^2^ < 0.001 for SNPs, we utilize the F statistic to assess the correlation between instrumental variables and exposure factors. Only when the F statistic exceeds 10, it is considered that there is no bias induced by weak instrumental variables[37].

### Statistical analyses

The exposure and outcome SNPs were extracted from the GWAS database, with the exposed SNPs serving as instrumental variables. Harmonization was then processed to make the effect alleles of the exposure and outcome SNPs coincide and rule out SNPs with incompatible alleles or being palindromic with intermediate effect allele frequency. Specifically, four steps were followed: (1) Independent genetic instrumental variables were obtained from the exposure database based on the aforementioned SNP selection method. (2) Proxied SNPs would be found for missed SNPs. (3) Significantly associated SNPs with outcomes were discarded. (4) Fuzzy and palindromic SNPs were excluded as well. Following the completion of normalization, MR Analysis was conducted using reverse variance weighted (IVW) estimates as primary MR effect estimates, reported as odds ratios (OR) along with 95% confidence intervals (95% CI)[38]. Two additional methods estimating causal effects included weighted median and MR-Egger. These three methods are widely recognized providing robust analysis results in Mendelian randomization studies[39]. For application of the weighted median method, at least 50% of SNP instruments must meet validity criteria[40]. The suitability of MR-Egger lies in its ability to detect potential violations of standard instrumental variable assumptions and provide effect estimates unaffected by such violations[35].

The MR analysis was conducted using R software (version 4.3.1), specifically the TwoSample MR package (version 0.5.7) and RadialMR package (version 1.0).

## Results

### Biochemical markers of iron status have no causal effect on thyroid function

We successfully extracted 19 SNPs representing biochemical markers of iron status from serum iron, ferritin, transferrin, and transferrin saturation data. In our study, the F statistics of iron status range from 41.59 to 331.74, larger than the conventional value of 10, indicating that the selected instrumentals had a strong potential to predict iron status.

We employed IVW, MR-Egger regression, and weighted median methods to assess the causal association between iron status and thyroid dysfunction, with IVW serving as the primary method for significance testing. The IVW results indicated that all exposure factors and outcome characteristics in the IVW test had *p-values* > 0.05, suggesting no significant causal relationship of biochemical markers of iron status on thyroid dysfunction was observed (Table 2).

**Table 2.**
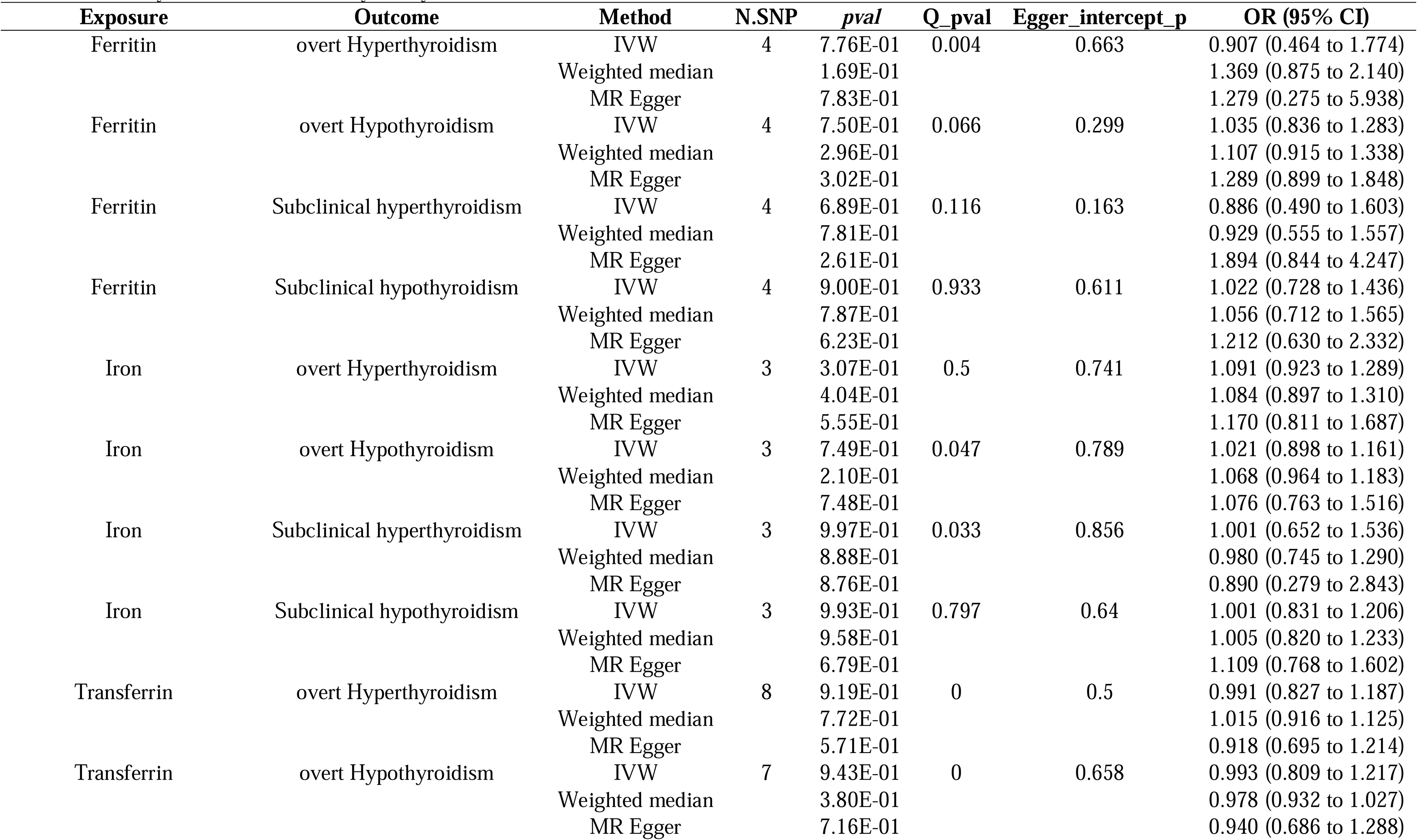
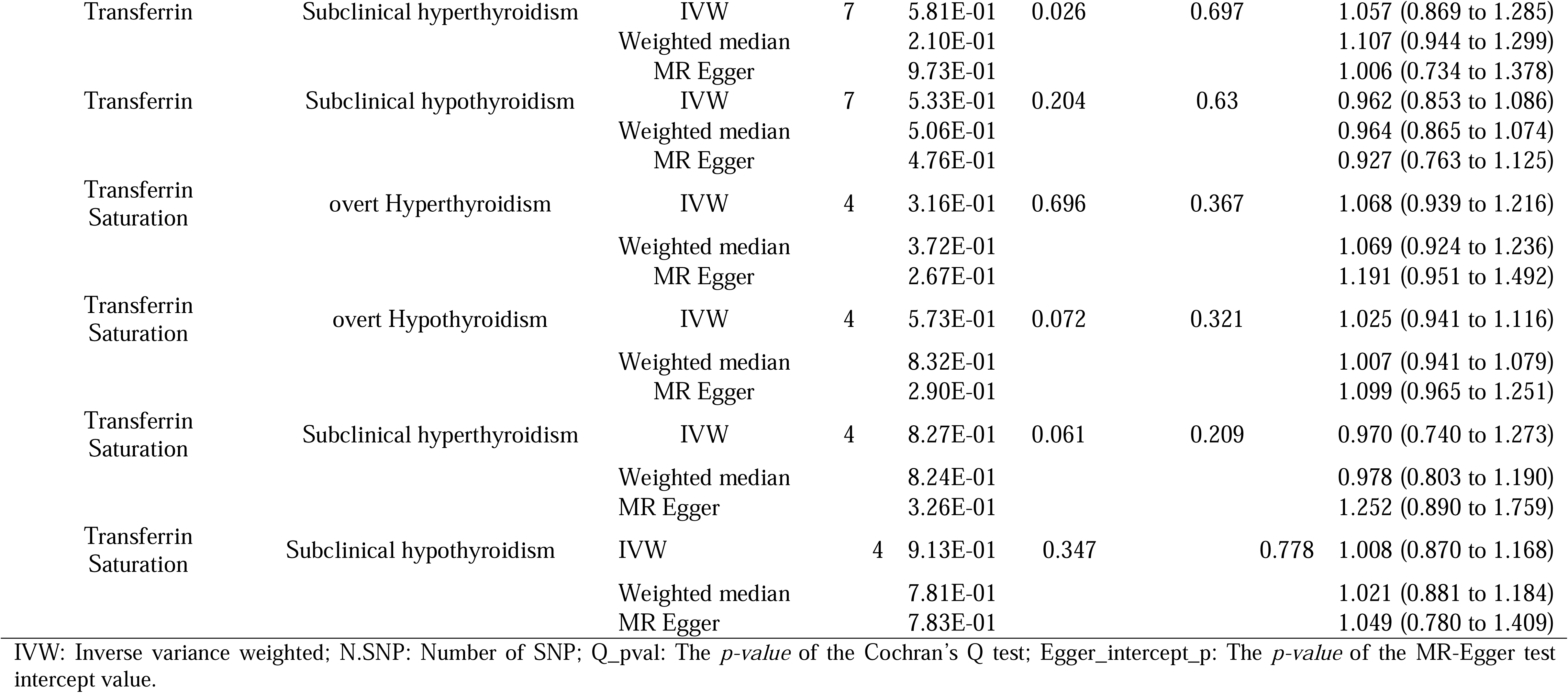
MR analysis of iron status and thyroid dysfunction.

### Biochemical markers of iron status have a causal effect on IDA

To verify the causal relationship between iron status biomarkers and IDA, we conducted two-sample MR analyses for serum iron, ferritin, transferrin saturation, and transferrin in relation to IDA. The instrumental variables employed were derived from the aforementioned biochemical markers of serum iron status.

We employed IVW, MR-Egger regression, and the weighted median method to assess the causal relationship between iron status and IDA. IVW was primarily used as the method for significance detection. The results from IVW indicated that serum iron (OR = 0.760, 95%CI = 0.651-0.888, *p* < 0.001), serum ferritin (OR = 0.518, 95%CI = 0.338-0.795, *p* = 0.003), and serum transferrin saturation (OR = 0.810, 95% CI = 0.718-0.915, *p* < 0.001) were causally associated with a reduced risk of IDA (Figure2). However, no significant causal relationship was observed of serum transferrin (OR = 1.080,95 % CI=0.986 –1 .182, *p* = 0.096) on IDA (shown in Fig. 2). Similar risk estimates were obtained using MR-Egger regression and the weighted median methods (shown in Fig. 2). The consistency among these 3 MR models enhances the reliability of the protective effect of these three markers of serum iron status in relation to IDA (shown in Supplementary Fig. S1)[41]. To thoroughly investigate any potential bias in this MR study, a sensitivity analysis was conducted using a complementary approach. The Cochran’s Q test revealed no heterogeneity, and there was no significant intercept, suggesting that pleiotropy was not observed (shown in Fig. 2). Furthermore, the Steiger Test found no reverse causal effect.

**Fig. 2.**
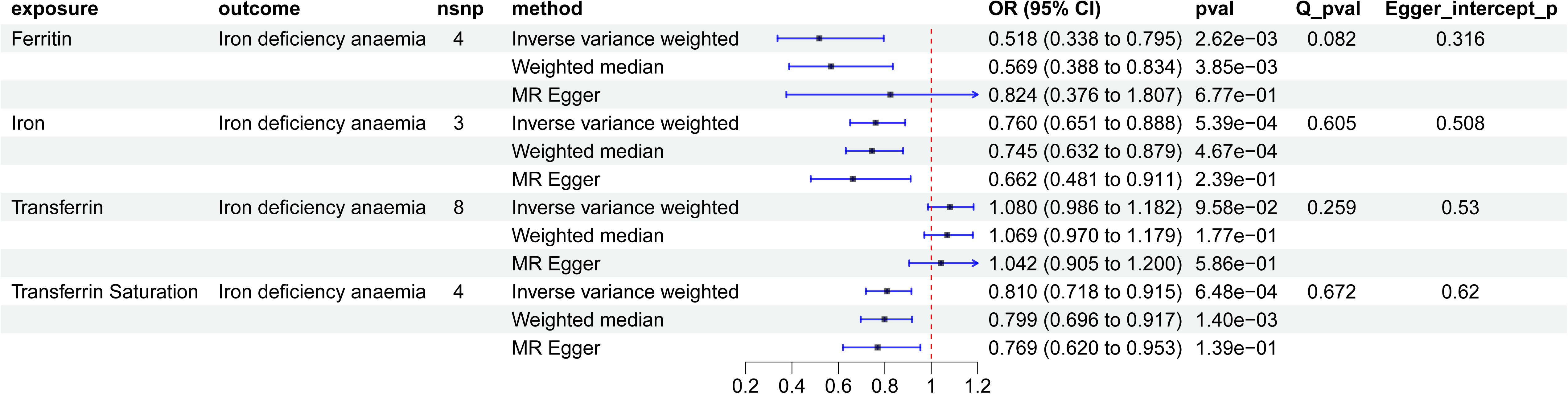
Forest plot of MR analysis of serum iron status and IDA.

The details of SNPs are provided in Supplementary Table S1[41]. Leave-one analysis was employed to validate the impact of each SNP on the overall causal estimate. As shown in Supplementary Figure S2[41], after excluding a specific SNP, the remaining SNPs exhibited no meta-effect exceeding 0 for the three serum iron status markers with significant causal relationships, indicating consistent and reliable results. Based on the aforementioned analysis, there was no violation observed in terms of the causal effect of serum iron, ferritin, and transferrin saturation on IDA.

### Overt hypothyroidism has a causal effect on IDA

A total of 106 SNPs associated with thyroid dysfunction were successfully identified from the datasets of subclinical and overt thyroid dysfunction. The instrumental variables derived from these SNPs exhibited F statistics ranging from 41.59 to 83.82, all surpassing the mean value of 10 and demonstrating strong instrumentality.

The result of IVW indicated strong evidence that overt hypothyroidism had a causal relationship with the increased risk of IDA (OR = 1.101, 95%CI = 1.048-1.157, *p* < 0.001), however, no significant association was found between other thyroid dysfunction on IDA (*p* > 0.05) (shown in Fig. 3). Meanwhile, similar risk estimates were gained using the MR-Egger regression (OR = 1.092, 95% CI = 0.981-1.215, *p* = 0.112) and weighted median approaches (OR = 1.19, 95% CI = 1.036-1.210, *p* = 0.005). The concordance of the 3 MR models enhanced the reliability of the risk role of overt hypothyroidism in the issue of IDA (shown in Supplementary Fig. 4a)[41]. Further, we conducted the MR-PRESSO and Radial MR analysis. No outlier was identified with MR-PRESSO but two outliers (rs12379417, rs4409785) were detected with Radial MR (shown in Supplementary Fig. 4b). We manually removed these outliers and repeated IVW analysis, and the causality remained (OR = 1.090, 95% CI =1.037–1.147, *p* < 0.001, shown in Supplementary Fig. 4c and d). Figure 3 also shows that the Cochran Q test found no significant heterogeneity and no evidence of significant intercept, that is, no pleiotropy was observed. In addition, the Steiger Test found no reverse causal effect.

**Fig. 3.**
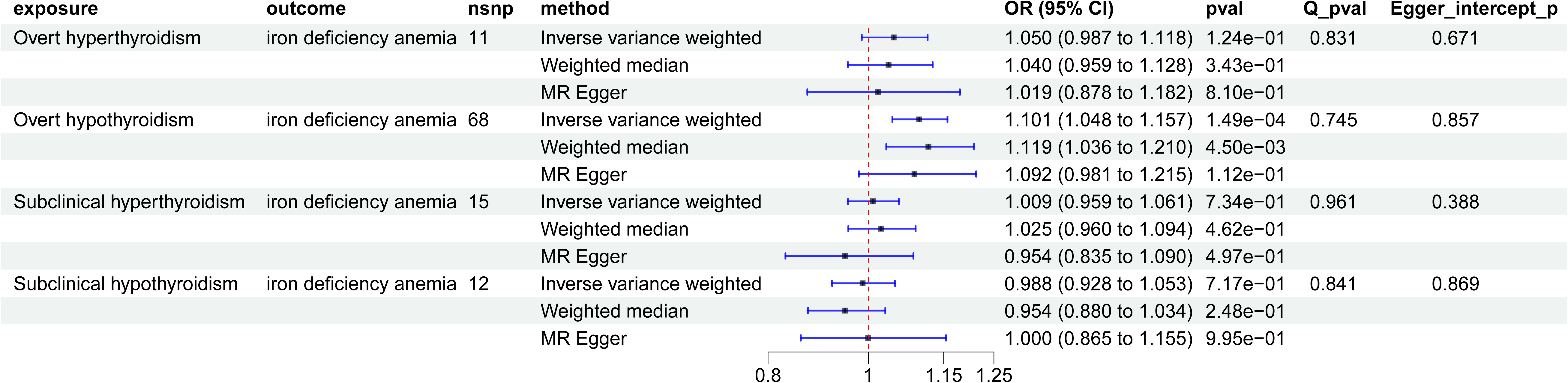
Forest plot of MR Analysis of thyroid dysfunction and IDA.

**Fig. 4.**
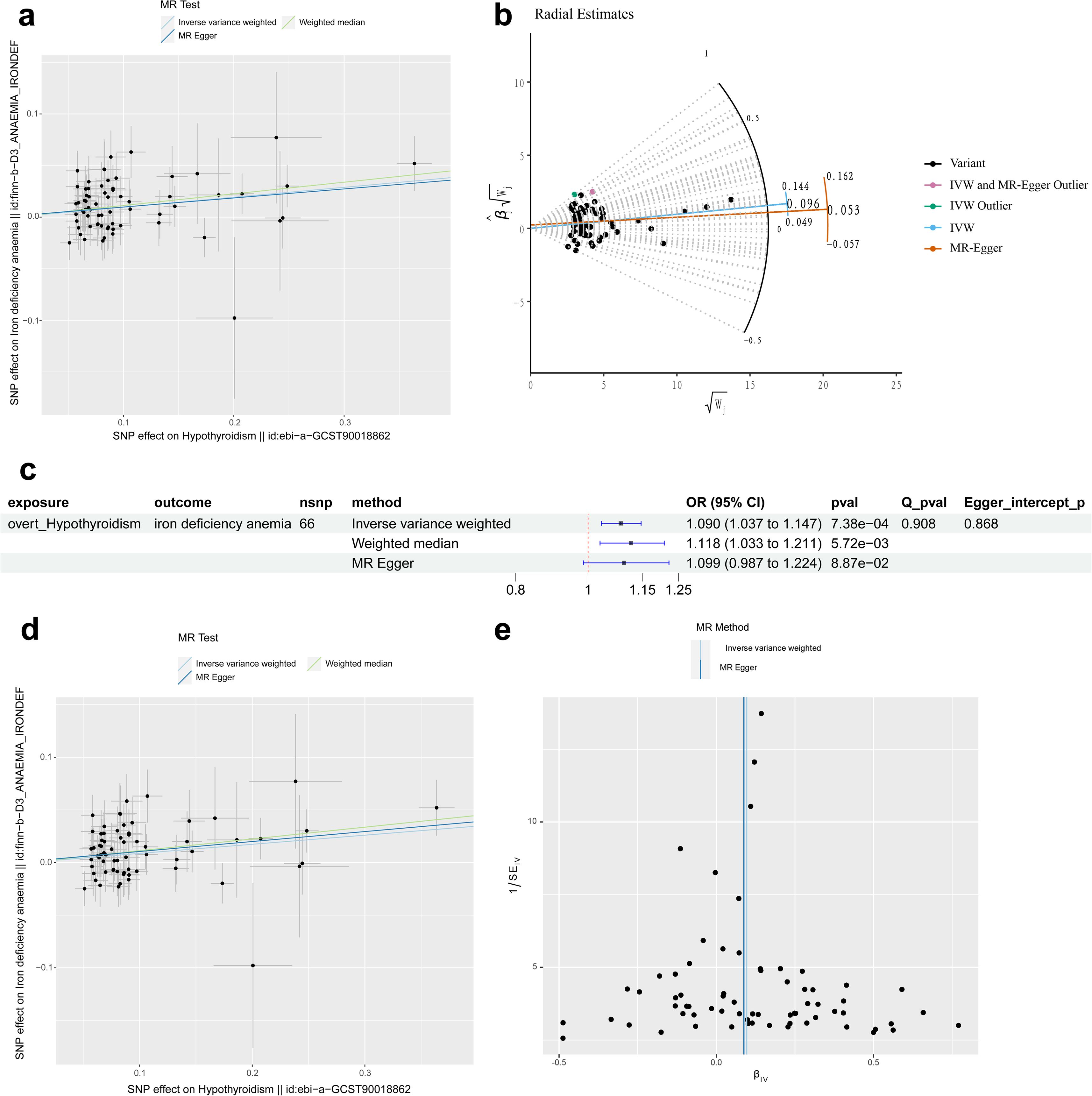
Combination of MR analysis of overt hypothyroidism and IDA Notes: (a) Scatter plot with no outliers removed; (b) Radial estimates; (c) Forest plot with outliers removed; (d) Scatter plot with outliers removed; (e) Funnel plot with outliers removed.

Details of SNPs are recorded in Supplementary Table S2[41]. The funnel plot is shown in Supplementary Figure 4E. The finding suggests that overt hypothyroidism does not violate the causal effect on IDA.

### Replicated analyses

We conducted a replicated MR analysis using another overt hypothyroidism data from the GWAS Catalog and the IDA data from the UK Biobank (Table 1). The IVW method revealed a persistent causal association between overt hypothyroidism and an increased risk of IDA (OR = 1.023, 95% CI = 1.011-1.035, *p* < 0.001). Consistent with this finding, both MR-Egger (OR = 1.031, 95% CI = 1.004-1.058, *p* =0.024) and the weighted median method yielded similar results (OR = 1.022, 95% CI = 1.003-1.040, *p* = 0.021) (shown in Supplementary Fig. S3a)[41]. The Cochran Q test and MR-Egger polymorphism test results are shown in Supplementary Figure S3a as well. The scatter plot and funnel plot are shown in Supplementary Figure S3b and d respectively. Furthermore, the Steiger Test did not reveal any evidence of reverse causation.

## Discussion

This is the first study to employ a two-sample MR approach, utilizing large-scale GWAS datasets, in order to investigate the causal relationship between human biochemical markers of iron status and thyroid dysfunction, as well as between thyroid dysfunction and IDA. In essence, our findings reveal that there is no significant evidence supporting a causal link between the four biochemical markers of iron status (serum iron, ferritin, transferrin, and transferrin saturation) and thyroid dysfunction based on IVW’s primary estimate. However, when employing the same method to analyze the relationship between thyroid dysfunction and IDA, we discovered that overt hypothyroidism exhibits a causal effect on IDA (OR = 1.101; 95% CI = 1.048-1.157; *p* < 0.001). The robustness of this result was further confirmed through secondary analysis using additional GWAS data pertaining to the causal effect between hypothyroidism and IDA. Furthermore, we also validated the causal relationship between biochemical markers of iron and IDA. Although this association may appear evident, our MR approach substantiates it from a SNP perspective while also demonstrating the reliability of this methodology.

Thyroid dysfunction, whether significant or subclinical, is a prevalent endocrine disorder affecting approximately 5-10% of the general population. Notably, hypothyroidism exhibits a higher prevalence compared to hyperthyroidism[42]. Several studies have indicated that thyroid function is dependent on iron status, as iron deficiency reduces thyroid peroxidase activity[14–16]. Efficient synthesis of thyroid hormones requires adequate iron status, as TPO only exhibits activity on the apical surface of thyroid cells after binding with the prosthetic heme group[11]. Observational studies have also reported a higher prevalence of iron deficiency and lower serum iron concentrations in patients with subclinical hypothyroidism or Hashimoto’s thyroiditis compared to healthy controls[43, 44]. However, our MR analysis did not reveal a direct causal relationship between iron status biomarkers and thyroid dysfunction (both dominant and subclinical). This may be attributed to the existence of undiscovered mechanisms and pathways linking the two variables. Notably, some studies have also reported no significant association between serum/plasma iron concentrations or ferritin levels and hypothyroidism[22, 45].

The development of IDA is attributed to an imbalance in the body’s iron absorption and excretion, resulting in a reduction of iron content. Insufficient intake, excessive demand, excessive loss, as well as disorders in absorption and utilization can all contribute to the occurrence of IDA[46–48]. IDA not only impacts quality of life but also poses potential risks to brain development and cognitive function [49]. Thyroid disease and IDA are significant clinical challenges due to their high prevalence and close association[50]. In some cases, iron deficiency anemia can serve as the initial indicator for diagnosing hypothyroidism, commonly referred to as the hematological manifestation of hypothyroidism[51]. The association between hypothyroidism and IDA in early pregnancy was demonstrated in an observational study[52]. Additionally, a study conducted in India by Das et al.[53] revealed that small cell anemia ranked as the second most prevalent type of anemia among individuals with hypothyroidism, following normal cell normal pigmentary anemia. An initial animal study[13] demonstrated that supplementation of T3 in hypothyroid rats restored gastrointestinal iron absorption, while treatment with T4 for 1 year in women with subclinical hypothyroidism and iron deficiency was associated with a reduced incidence of anemia and slightly elevated levels of ferritin, iron, and Hb[44]. Furthermore, a recent study indicated that the utilization of a novel liquid L-T4 preparation can be concomitantly administered with iron salts without compromising absorption[45]. Therefore, elucidating the precise relationship between hypothyroidism and IDA plays a pivotal role in enhancing the therapeutic efficacy of IDA or combination therapy for hypothyroidism. However, no other study has been able to establish causation or correlation with sufficient empirical evidence. The existing inconsistent conclusions may arise from various confounding factors, such as disparate study designs and diverse populations. In our investigation, we employed Mendelian randomization analysis as a methodological approach. This technique utilizes genetic tools to infer potential causality, mitigates bias resulting from confounding variables, and estimates assumed causality under varying conditions. Furthermore, we replicated the analysis using an independent GWAS dataset. We also performed sensitivity analyses and identified potential risk factors to ensure compliance with assumptions. Consequently, our findings provide robust evidence supporting the causal effect of hypothyroidism on IDA, but it’s important to note that IVs estimates may provide causal effects only under certain assumptions.

Currently, there is no conclusive understanding of the underlying mechanism by which hypothyroidism affects IDA. Several potential mechanisms have been proposed. A previous study using MR analysis [54] indicated a causal relationship between thyroid function and pernicious anemia. It has been speculated that this association may be attributed to autoimmune comorbidities accompanying hypothyroidism, such as autoimmune gastritis, leading to impaired absorption of vitamin B12 in the stomach. Similarly, the causal link between overt hypothyroidism and iron deficiency anemia can also be explained through this inference; certain autoimmune gastrointestinal disorders like celiac disease or autoimmune gastritis could hinder iron and other nutrient absorption in the gastrointestinal tract, consequently depleting body iron stores and resulting in iron deficiency anemia[55, 56]. However, these hypotheses regarding the mechanistic pathway are speculative at present, necessitating further relevant studies for confirmation.

To the best of our knowledge, this study represents the first MR analysis exploring the causal relationship between biochemical markers of iron status and thyroid dysfunction, as well as thyroid dysfunction and IDA. Our research possesses several key strengths. Firstly, both exposure factors and outcome signature genes instrumental variables were derived from European populations in order to mitigate potential confounding due to population stratification. Secondly, the selected GWAS dataset encompasses a large sample size with millions of detected SNPs, thereby substantially enhancing statistical power. Most importantly, we conducted repeated analyses and employed multiple supplementary methods to ensure result stability and reinforce estimate robustness. Our findings suggest that when treating IDA with iron supplementation, clinicians should consider assessing for hypothyroidism if the treatment response is unsatisfactory. In cases where hypothyroidism is present, it is recommended to address or control hypothyroidism symptoms prior to focusing on IDA treatment in order to potentially enhance therapeutic efficacy. Conversely, patients diagnosed with hypothyroidism should be vigilant for possible comorbidities such as IDA.

The limitations of this study should also be acknowledged in this manuscript. Firstly, our findings are specific to the European population, and further research is needed to generalize these conclusions to other populations. Secondly, additional validation is required to establish the underlying mechanism through which overt hypothyroidism increases the risk of IDA in European populations. Lastly, due to the utilization of summary-level statistics only, stratified analysis was not feasible.

## Conclusion

In our MR study, an upregulation of the gene associated with hypothyroidism was found to be positively correlated with an elevation in IDA prevalence within the European population. These findings have important clinical implications, suggesting that healthcare providers should consider coexisting hypothyroidism and IDA when designing treatment strategies. In cases where necessary, a combined therapeutic approach may be warranted to optimize treatment efficacy.

## Statements

## Data Availability

All data produced in the present study are available upon reasonable request to the authors.

https://figshare.com/

## Acknowledgement

The authors would like to thank all the genetics consortiums for making the GWAS summary data publicly available.

## Statement of Ethics

In all corresponding original studies, informed consent was obtained from all participants, and since only summary-level statistics were employed in our analysis, no additional ethical approval was required.

## Author Contributions

XH and TG designed the study. XH, TG, and YW conducted research. XH and TG analyzed the data. XH, TG, YW, QX and JD wrote this paper. All authors contributed to the article and approved the submitted version.

## Data Availability Statement

All data generated or analyzed during this study are included in this published article or in the data repositories listed in “References”, further inquiries can be directed to the corresponding author.

## Supplementary figures

Fig. S1. Scatter plot of MR analysis of serum iron status and IDA

Notes: The exposure factor is (a) Iron; (b) Ferritin; (c) Transferritin saturation and (d) Transferritin.

Fig. S2. Leave-one-out analysis of serum iron status and IDA

Notes: The exposure factor is (a) Iron; (b) Ferritin; (c) Transferritin saturation and (d)

Fig. S3. Overt hypothyroidism and IDA replicated MR analysis results

Notes: (a) Forest plot; (b) Scatter plot; (c) Funnel plot.

