## Supplementary figures and images for "Iron status, thyroid dysfunction and iron deficiency anemia: a two-sample Mendelian randomization study"

### Supplemental Figure S1

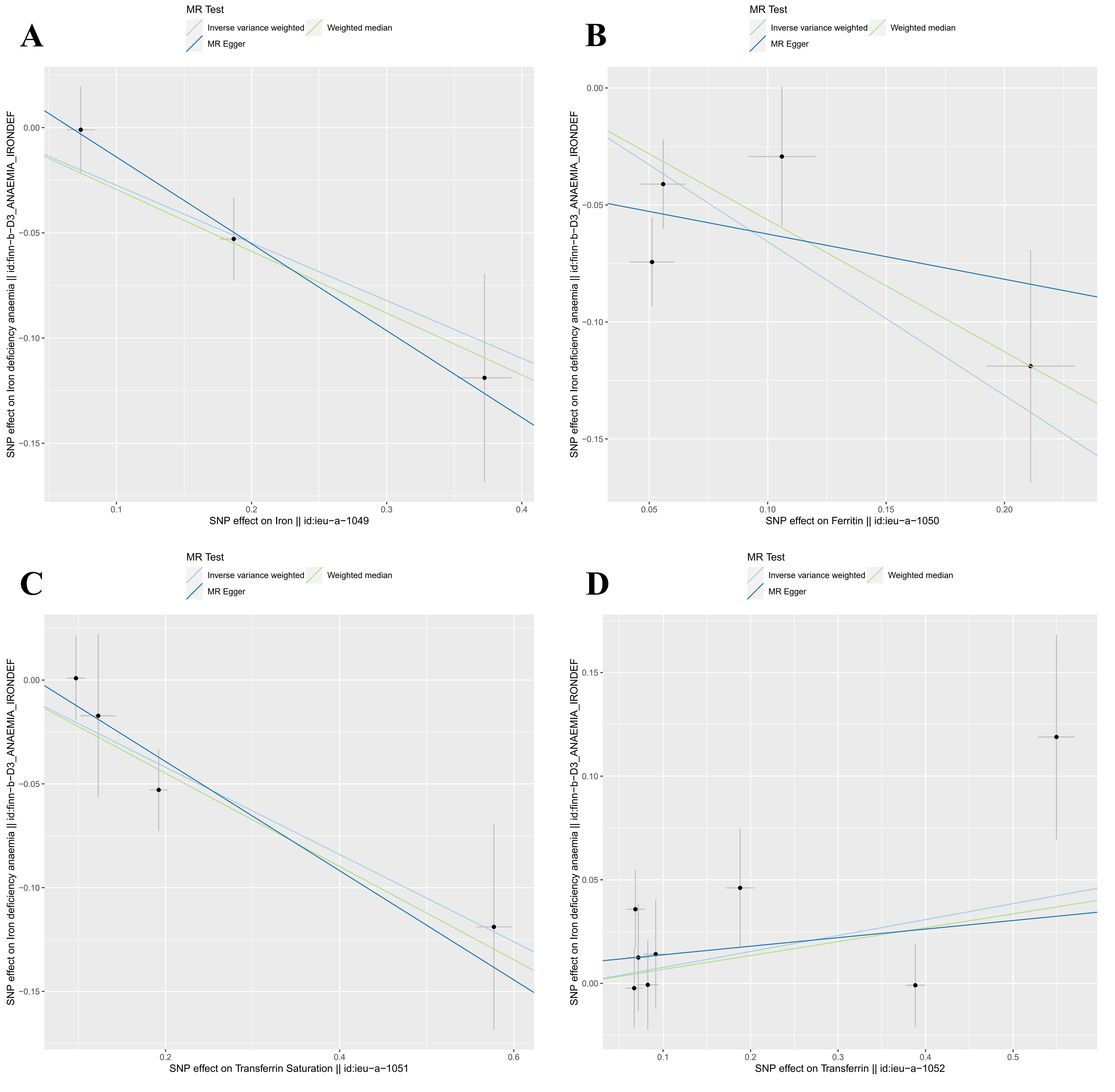

### Supplemental Figure S2

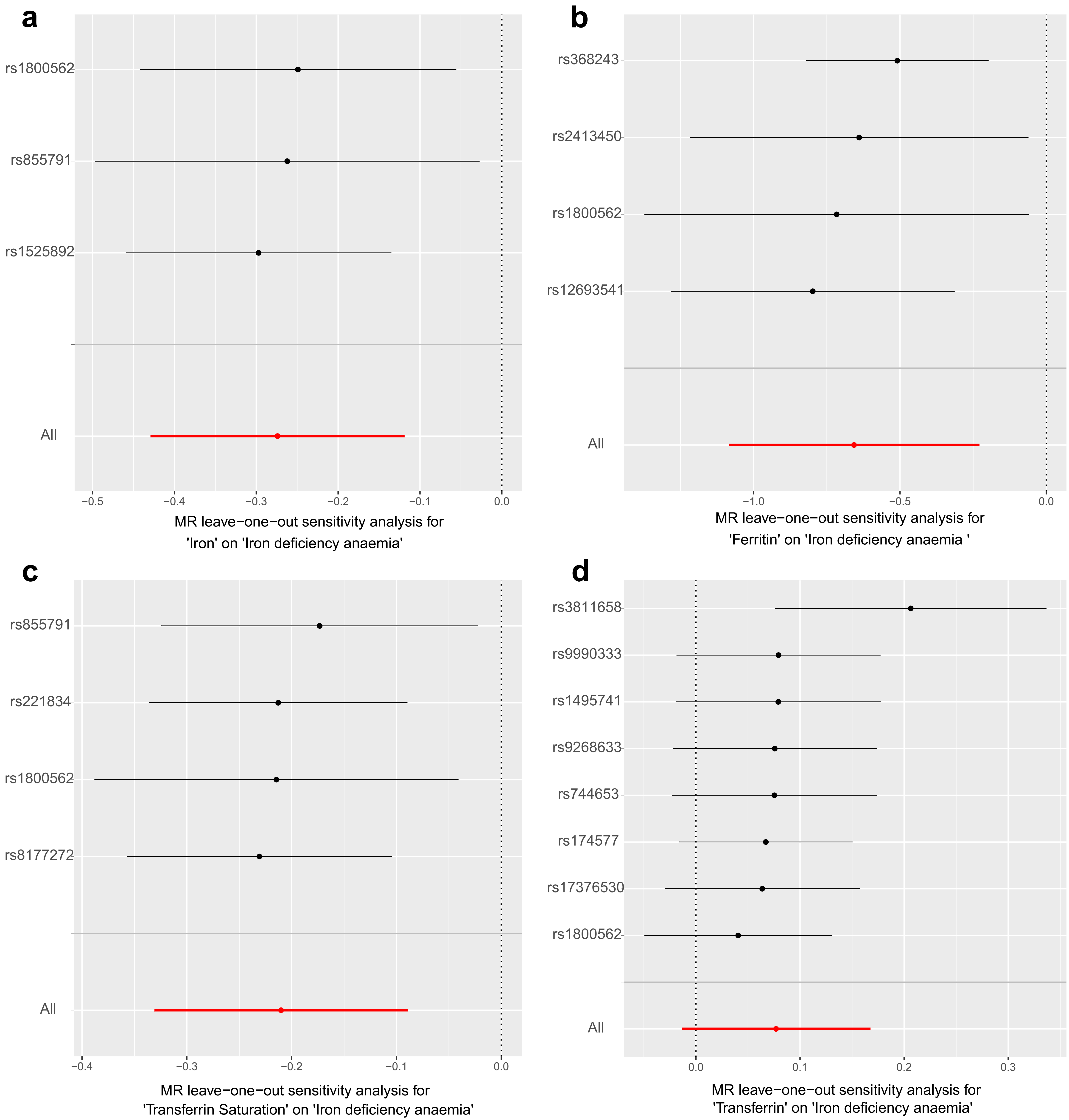
