## Supplemental Figure S3 for "Iron status, thyroid dysfunction and iron deficiency anemia: a two-sample Mendelian randomization study"

**a**

| exposure | outcome | nsnp | method | OR (95% CI) | pval | Q_pval | Egger_intercept_p |
|---|---|---|---|---|---|---|---|
| Hypothyroidism | Iron deficiency anaemia | 122 | Inverse variance weighted | 1.023 (1.011 to 1.035) | 1.09e−04 | 0.127 | 0.542 |
| | | | MR Egger | 1.031 (1.004 to 1.058) | 2.44e−02 | | |
| | | | Weighted median | 1.022 (1.003 to 1.040) | 2.09e−02 | | |

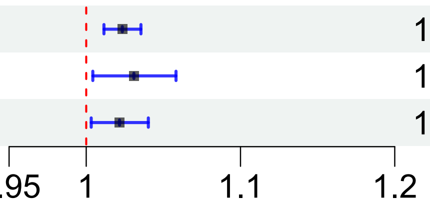


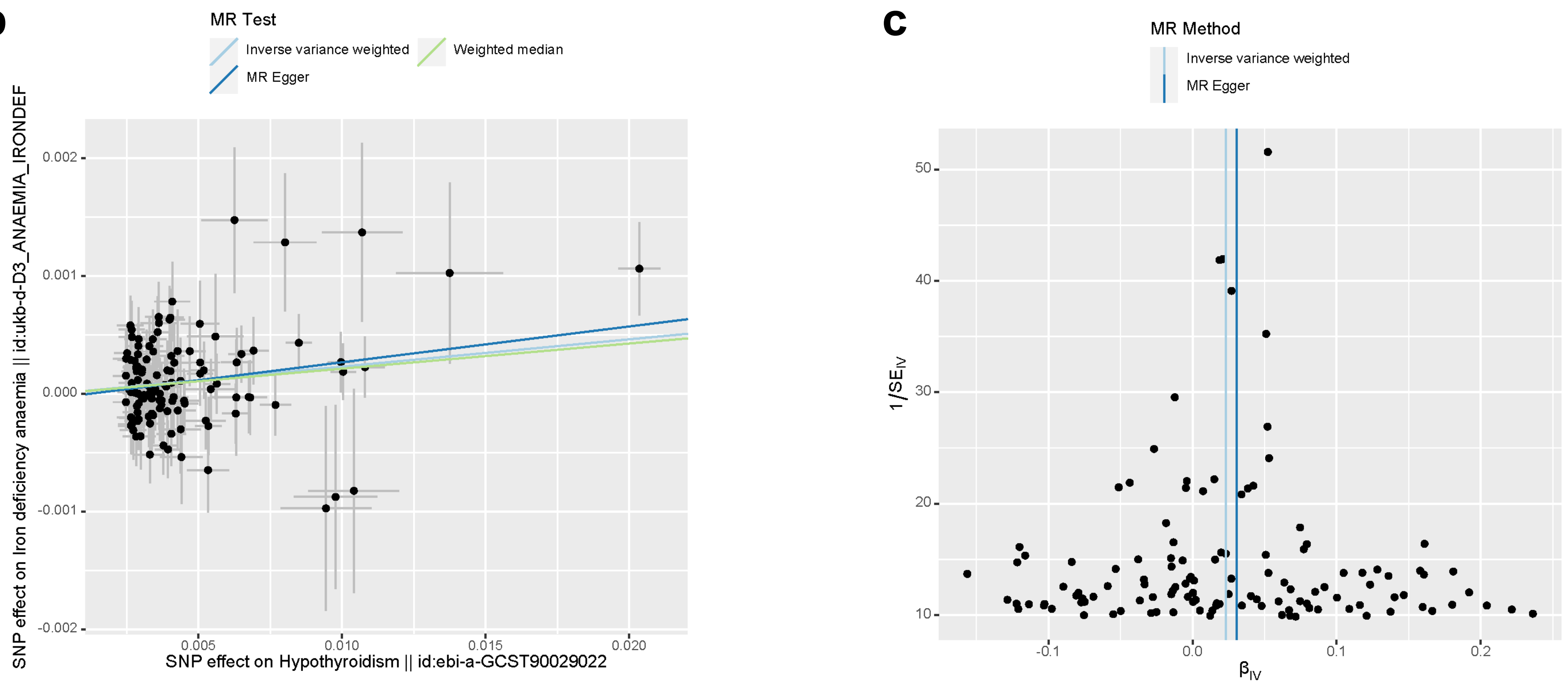
