## Supplemental Tables for "Iron status, thyroid dysfunction and iron deficiency anemia: a two-sample Mendelian randomization study"

**Table S1:** Details of the instrument variables for serum biochemical markers of iron status included in MR analysis.

| **Exposure** | **SNP** | **effect allele** | **other allele** | **beta** | **eaf** | **se** | **pval** |
| --- | --- | --- | --- | --- | --- | --- | --- |
| Iron | rs1525892 | A | G | 0.0736 | 0.347900003 | 0.0104 | 1.65E-12 |
| Iron | rs1800562 | A | G | 0.3724 | 0.042739999 | 0.02 | 3.96E-77 |
| Iron | rs855791 | G | A | 0.1868 | 0.387700021 | 0.0101 | 4.31E-77 |
| Ferritin | rs12693541 | T | C | -0.106 | 0.146099985 | 0.014 | 4.18E-14 |
| Ferritin | rs1800562 | A | G | 0.211 | 0.042739999 | 0.0187 | 1.42E-29 |
| Ferritin | rs368243 | C | T | -0.0512 | 0.451300025 | 0.0093 | 3.80E-08 |
| Ferritin | rs2413450 | C | T | 0.0559 | 0.419499993 | 0.0095 | 3.57E-09 |
| Transferrin Saturation | rs8177272 | A | G | -0.097 | 0.346899986 | 0.0106 | 5.52E-20 |
| Transferrin Saturation | rs1800562 | A | G | 0.5772 | 0.042739999 | 0.0203 | 1.52E-178 |
| Transferrin Saturation | rs221834 | G | C | 0.1226 | 0.053679999 | 0.0205 | 2.38E-09 |
| Transferrin Saturation | rs855791 | G | A | 0.1921 | 0.387700021 | 0.0101 | 3.50E-80 |
| Transferrin | rs744653 | T | C | 0.0916 | 0.156099975 | 0.0144 | 2.00E-10 |
| Transferrin | rs3811658 | T | C | 0.3883 | 0.347900003 | 0.0109 | 1.00E-200 |
| Transferrin | rs9990333 | T | C | -0.067 | 0.471199989 | 0.0101 | 3.01E-11 |
| Transferrin | rs17376530 | T | C | -0.1881 | 0.082500003 | 0.0165 | 5.43E-30 |
| Transferrin | rs1800562 | A | G | -0.5496 | 0.042739999 | 0.0208 | 1.26E-153 |
| Transferrin | rs9268633 | G | A | 0.0717 | 0.185899973 | 0.0128 | 2.31E-08 |
| Transferrin | rs1495741 | A | G | 0.0825 | 0.243499994 | 0.0122 | 1.57E-11 |
| Transferrin | rs174577 | A | C | 0.0684 | 0.360799998 | 0.0107 | 1.90E-10 |

**Table S2:** Details of the instrument variables for overt hypothyroidism included in MR analysis.

| **Exposure** | **SNP** | **effect allele** | **other allele** | **beta** | **eaf** | **se** | **pval** |
| --- | --- | --- | --- | --- | --- | --- | --- |
| Overt hypothyroidism | rs10075764 | G | A | -0.057 | 0.302413 | 0.0104 | 4.26E-08 |
|  | rs10126000 | A | C | -0.0683 | 0.688692 | 0.0104 | 5.13E-11 |
|  | rs10424978 | A | C | -0.0775 | 0.624328 | 0.0102 | 2.77E-14 |
|  | rs1079418 | G | A | -0.0657 | 0.262374 | 0.011 | 2.14E-09 |
|  | rs10917477 | G | A | 0.064 | 0.386901 | 0.01 | 1.75E-10 |
|  | rs11171710 | A | G | -0.0698 | 0.443916 | 0.01 | 3.19E-12 |
|  | rs11406335 | TG | T | -0.057 | 0.417169 | 0.0103 | 3.44E-08 |
|  | rs114285740 | C | G | 0.1669 | 0.0273776 | 0.0301 | 3.06E-08 |
|  | rs11675342 | T | C | 0.0906 | 0.387729 | 0.01 | 1.40E-19 |
|  | rs11875260 | G | A | 0.0751 | 0.172925 | 0.0135 | 2.54E-08 |
|  | rs12117927 | A | C | 0.0627 | 0.476541 | 0.0105 | 2.29E-09 |
|  | rs12582330 | T | G | -0.061 | 0.624365 | 0.0109 | 2.05E-08 |
|  | rs12593201 | A | G | 0.0905 | 0.325432 | 0.0112 | 7.69E-16 |
|  | rs12984428 | A | G | -0.0659 | 0.35604 | 0.0102 | 1.11E-10 |
|  | rs13090803 | T | G | 0.0829 | 0.190624 | 0.0128 | 9.00E-11 |
|  | rs13109179 | A | G | 0.0647 | 0.461895 | 0.01 | 9.42E-11 |
|  | rs1364450 | C | A | 0.0886 | 0.126235 | 0.0139 | 1.97E-10 |
|  | rs142997491 | G | A | 0.2385 | 0.0135637 | 0.0412 | 7.02E-09 |
|  | rs1432806 | G | A | 0.0583 | 0.338619 | 0.0105 | 2.89E-08 |
|  | rs1479565 | A | G | 0.0975 | 0.498866 | 0.0101 | 7.53E-22 |
|  | rs1534430 | T | C | -0.086 | 0.428394 | 0.0101 | 1.44E-17 |
|  | rs187707293 | A | T | 0.2419 | 0.0133864 | 0.044 | 3.99E-08 |
|  | rs2111485 | G | A | 0.0813 | 0.479023 | 0.0102 | 1.43E-15 |
|  | rs2114702 | A | T | 0.07 | 0.26671 | 0.0111 | 3.00E-10 |
|  | rs2234167 | A | G | 0.0825 | 0.102013 | 0.015 | 3.75E-08 |
|  | rs2247314 | C | T | -0.086 | 0.376495 | 0.0104 | 1.06E-16 |
|  | rs229528 | T | C | 0.0903 | 0.496016 | 0.01 | 2.31E-19 |
|  | rs2412976 | G | C | 0.0637 | 0.478888 | 0.0103 | 5.47E-10 |
|  | rs2445608 | A | G | -0.0593 | 0.430366 | 0.0101 | 3.79E-09 |
|  | rs244685 | G | T | -0.0858 | 0.79915 | 0.0132 | 7.06E-11 |
|  | rs2921053 | C | G | -0.0599 | 0.589616 | 0.0101 | 3.36E-09 |
|  | rs2988277 | T | C | 0.0593 | 0.287022 | 0.0106 | 2.49E-08 |
|  | rs307558 | A | G | -0.0688 | 0.742625 | 0.0119 | 8.01E-09 |
|  | rs3087243 | A | G | -0.1466 | 0.386088 | 0.0102 | 4.77E-47 |
|  | rs3118469 | T | A | 0.0803 | 0.295174 | 0.0106 | 3.82E-14 |
|  | rs3184504 | C | T | -0.1734 | 0.668513 | 0.0102 | 7.50E-65 |
|  | rs34536443 | C | G | -0.1863 | 0.0440279 | 0.0263 | 1.46E-12 |
|  | rs3775291 | T | C | -0.0649 | 0.287788 | 0.0108 | 1.65E-09 |
|  | rs434294 | G | A | -0.0683 | 0.322802 | 0.0109 | 3.36E-10 |
|  | rs4529854 | T | C | -0.0768 | 0.723438 | 0.0107 | 6.56E-13 |
|  | rs4835534 | C | T | -0.1421 | 0.156356 | 0.0132 | 7.06E-27 |
|  | rs5912815 | G | T | -0.0511 | 0.576761 | 0.0084 | 1.05E-09 |
|  | rs61759532 | T | C | 0.0905 | 0.188941 | 0.0122 | 1.42E-13 |
|  | rs61877856 | T | C | -0.0658 | 0.197495 | 0.0115 | 1.14E-08 |
|  | rs6679677 | A | C | 0.3637 | 0.108444 | 0.0159 | 2.39E-115 |
|  | rs6908626 | T | G | 0.1441 | 0.17075 | 0.0141 | 2.04E-24 |
|  | rs7030280 | T | C | 0.2075 | 0.745664 | 0.0108 | 1.02E-82 |
|  | rs71508903 | T | C | 0.0934 | 0.210816 | 0.0125 | 9.34E-14 |
|  | rs7223956 | C | T | -0.0902 | 0.897005 | 0.0144 | 4.27E-10 |
|  | rs73192661 | T | C | -0.1061 | 0.428858 | 0.01 | 4.05E-26 |
|  | rs736374 | A | G | 0.0832 | 0.375103 | 0.0103 | 6.00E-16 |
|  | rs7441808 | G | A | 0.0766 | 0.21091 | 0.0111 | 5.17E-12 |
|  | rs7488011 | T | C | 0.1052 | 0.356485 | 0.0111 | 2.52E-21 |
|  | rs7574865 | G | T | -0.1321 | 0.74252 | 0.0117 | 1.67E-29 |
|  | rs7742626 | C | T | 0.0686 | 0.267343 | 0.0116 | 3.41E-09 |
|  | rs78765971 | G | GAC | 0.2444 | 0.146942 | 0.0162 | 1.68E-51 |
|  | rs79490353 | C | T | 0.2006 | 0.0241151 | 0.0349 | 8.82E-09 |
|  | rs7990020 | C | A | 0.0577 | 0.453482 | 0.0101 | 9.97E-09 |
|  | rs853305 | C | T | -0.0802 | 0.718894 | 0.0111 | 4.40E-13 |
|  | rs881858 | A | G | 0.0665 | 0.747899 | 0.0108 | 8.46E-10 |
|  | rs911760 | A | C | 0.0879 | 0.212143 | 0.0125 | 1.95E-12 |
|  | rs926103 | C | T | -0.0678 | 0.697985 | 0.0104 | 7.65E-11 |
|  | rs9264277 | C | T | -0.0862 | 0.645071 | 0.0111 | 9.03E-15 |
|  | rs9271365 | G | T | 0.2484 | 0.443466 | 0.0105 | 4.91E-123 |
|  | rs9277559 | C | T | -0.133 | 0.331026 | 0.012 | 1.86E-28 |
|  | rs9497965 | T | C | 0.0827 | 0.409035 | 0.0102 | 3.71E-16 |
|  | rs9511151 | A | G | -0.0976 | 0.279567 | 0.0106 | 3.08E-20 |
|  | rs9902341 | T | C | 0.0801 | 0.172257 | 0.0129 | 4.68E-10 |
